# An Explainable Machine Learning Framework for Predicting the Risk of Buprenorphine Treatment Discontinuation for Opioid Use Disorder

**DOI:** 10.1101/2023.11.02.23297982

**Authors:** Jabed Al Faysal, Md. Noor-E-Alam, Gary J. Young, Wei-Hsuan Lo-Ciganic, Amie J. Goodin, James L. Huang, Debbie L. Wilson, Tae Woo Park, Md Mahmudul Hasan

**Author notes:** **Corresponding Author:** Md Mahmudul Hasan, PhD, 1225 Center Drive, PO Box 100496, Gainesville, FL 32610.

## Abstract

**Objectives:** Buprenorphine is an effective evidence-based medication for opioid use disorder (OUD). Yet premature discontinuation undermines treatment effectiveness, increasing risk of mortality and overdose. We developed and evaluated a machine learning (ML) framework for predicting buprenorphine care discontinuity within 1-year following treatment initiation.

**Methods:** This retrospective study used United States 2018-2021 MarketScan commercial claims data of insured individuals aged 18-64 who initiated buprenorphine between July 2018 and December 2020 with no buprenorphine prescriptions in the previous six months. We measured buprenorphine prescription discontinuation gaps of ≥30 days within the first year of initiating treatment. We developed predictive models employing logistic regression, decision tree classifier, random forest, XGBoost, Adaboost, and random forest-XGBoost ensemble. We applied recursive feature elimination with cross-validation to reduce dimensionality and identify the most predictive features while maintaining model robustness. We focused on two distinct treatment stages: at the time of treatment initiation and one and three months after treatment initiation. We employed SHapley Additive exPlanations (SHAP) analysis that helped us explain the contributions of different features in predicting buprenorphine discontinuation. We stratified patients into risk subgroups based on their predicted likelihood of treatment discontinuation, dividing them into decile subgroups. Additionally, we used a calibration plot to analyze the reliability of the models.

**Results:** A total of 30,373 patients initiated buprenorphine and 14.98% (4,551) discontinued treatment. C-statistic varied between 0.56 and 0.76 for the first-stage models including patient-level demographic and clinical variables. Inclusion of proportion of days covered (PDC) measured at one-month and three-month following treatment initiation significantly increased the models’ discriminative power (C-statistics: 0.60 to 0.82). Random forest (C-statistics: 0.76, 0.79 and 0.82 with baseline predictors, one-month PDC and three-month PDC, respectively) outperformed other ML models in discriminative performance in all stages (C-statistics: 0.56 to 0.77). Most influential risk factors of discontinuation included early stage medication adherence, age, and initial days of supply.

**Conclusion:** ML algorithms demonstrated a good discriminative power in identifying patients at higher risk of buprenorphine care discontinuity. The proposed framework may help healthcare providers optimize treatment strategies and deliver targeted interventions to improve buprenorphine care continuity.

## 1. Introduction

Opioid use disorder (OUD) has emerged as a major public health crisis, affecting millions of people worldwide and imposing substantial social and economic burdens. Over 2.7 million individuals in the United States (US) struggle with OUD. Overdose deaths involving opioids quadrupled from approximately 21,000 in 2010 to over 80,000 in 2021 (NIH, 2023). In response, payers, policy makers, and healthcare systems have sought to expand access to treatment options for individuals with OUD.

Common treatment options for OUD include medications for opioid use disorder (MOUD) including methadone, buprenorphine, and naltrexone. Buprenorphine is an FDA approved evidence-based medication that offers reduced toxicity and greater outpatient accessibility compared to methadone (Walsh, 1995; Jaffe, 2003; Fiscella, 2019). It is proven to be effective in minimizing opioid cravings, overdose risk, and mortality risk for patients who continue the treatment for >6 months (Parran, 2010; Weiss, 2011; Fiellin, 2014). Despite its efficacy, the rates of discontinuation are higher compared to the individuals receiving methadone (Gryczynski, 2014; Mattick, 2014). Several studies reported that a significant proportion of individuals discontinued buprenorphine within the first six months of use, which substantially increases their likelihood for relapse, hospitalization, and even death (Samples, 2018; Shcherbakova, 2018).

Accordingly, identifying patients at risk of premature discontinuation of buprenorphine can help healthcare providers develop patient support systems that improve treatment retention rates (Saloner, 2023). Towards this goal, several studies have sought to identify patient risk factors for buprenorphine discontinuation using either conventional statistical approaches or machine learning procedures (Samples, 2018; Vakkalanka, 2022). These studies generally suggest that the risk for premature discontinuation is greater for patients who are under age 30, male, and of lower socio-economic status. Some research also suggests that those under the care of prescribers with a large number of patients for OUD treatment are at higher risk of treatment discontinuation (Hasan M. M., 2021). Medication dosage has also been shown to be relevant with lower initial dosage associated with a higher risk of discontinuation (Samples, 2018).

While existing research has helped identify certain risk factors for treatment discontinuation, most studies have not considered the predictive value of risk factors during different stages of the treatment process. The predictive value of a risk factor before treatment begins may be different than it is once treatment is initiated and additional information on the patient’s progress becomes available.

Our prior work developed a machine learning-based framework to predict buprenorphine treatment discontinuation in two distinct treatment stages: at the time of initiating treatment and during stabilization or early maintenance treatment phases (Hasan M. M., 2021). The use of two-staged setting has substantial clinical significance. First, prediction made at treatment initiation may help identify patients who might face challenges while starting the treatment and whether she/he is a good candidate for buprenorphine treatment. Then, the prediction made after one month and three months with information about a patient’s early medication adherence may help identify patients struggling to maintain the treatment, allowing for timely strategies to improve adherence and prevent premature discontinuation. However, this previous investigation was limited by a small sample that was drawn from a single state and by a relatively narrow set of clinical variables for prediction purposes.

Thus, building on our prior work, we aimed to develop a robust two-stage clinical decision support framework, utilizing machine learning and a large, nationally representative dataset to predict patients’ buprenorphine discontinuation. In addition, we aimed to incorporate model calibration that improves the predictive reliability of our ML framework and provides more detailed insights into the likelihood of patients’ buprenorphine discontinuation. We also aimed to introduce a risk stratification approach to categorize patients into subgroups with similar risks of buprenorphine treatment discontinuation.

## 2. Materials and Methods

### 2.1. Data Source

This study used the MarketScan commercial claims data from 2018 to 2021, which contains information on >43 million commercially insured patients and covers a wide range of healthcare services such as medical care received in inpatient and outpatient settings, prescription medications, and various medical tests used for diagnosis purposes (Butler, 2021). MarketScan provides longitudinal data and enables tracking patient-level information over time. This study was conducted in compliance with ethical standards and was approved by the University of Florida Institutional Review Board (IRB) under the reference number IRB202102917 with the requirement for written informed consent waived due to the nature of the data.

### 2.2. Study Design

Complying with TRIPOD guidelines (Moons, 2015), this prognostic predictive modeling study with retrospective cohort study design included patients with an OUD diagnosis (F11.1X and F11.2X) who initiated buprenorphine treatment during 2018-2021 and were 18–64 years old at treatment initiation and were not enrolled in Medicare Advantage plans. The date of the first buprenorphine claim (between 1 July 2018 and 31 December 2020) was set as the index date. We used national drug codes (NDCs) recorded in the prescription drug claims to identify these patients excluding those who filled buprenorphine only approved by the US Food and Drug Administration (FDA) for pain (intravenous or transdermal formulations) not OUD (Lo-Ciganic, 2016). Patients having a buprenorphine prescription for OUD in the 6-month pre-index period (i.e., baseline period) were also excluded from the study. We also excluded patients who had <2 buprenorphine fills, as it was more likely that these were prescribed for detoxification purposes rather than for maintenance treatment (Lo-Ciganic W. H., 2020; Lo-Ciganic, 2016). We restricted to patients who were continuously enrolled in a health insurance plan in the 6 months before and 12 months after the index date to ensure uninterrupted insurance coverage. Figure 1 represents the overall study design and Figure 2 illustrate the inclusion/exclusion criteria for the study cohort.

**Figure 1.**
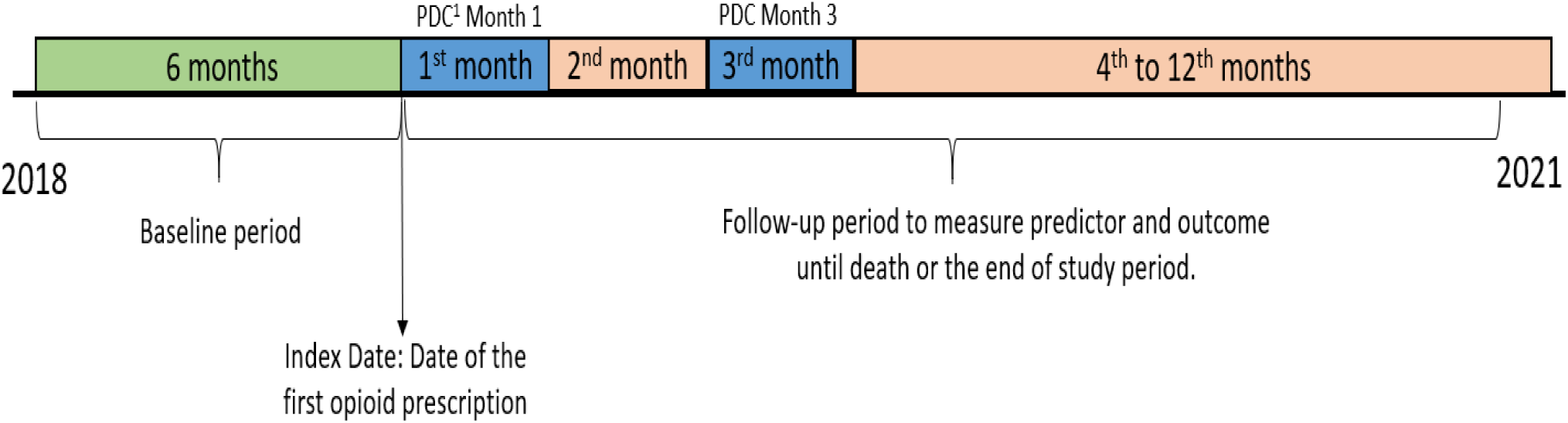
Overall Study Design Timeframe^1^

**Figure 2.**
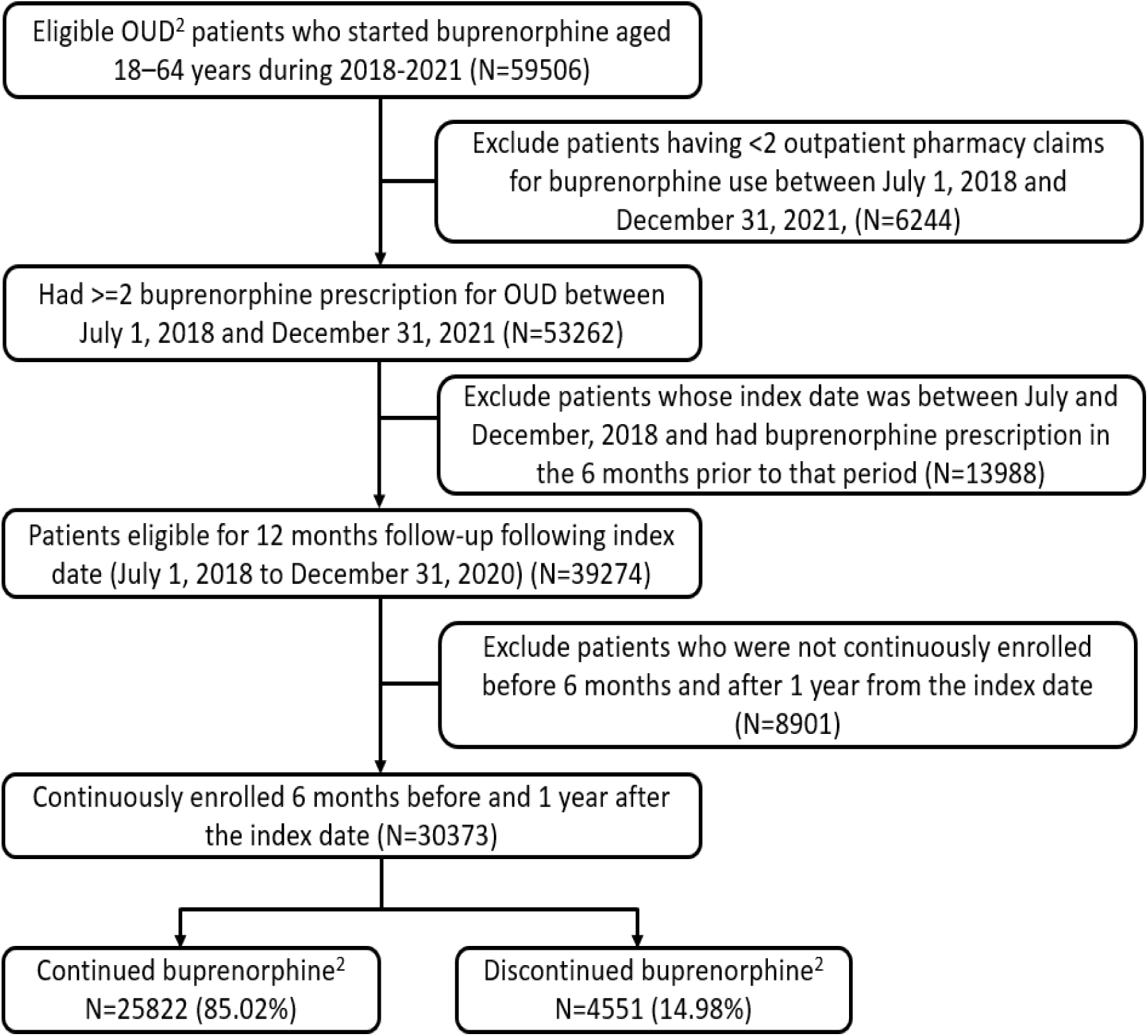
Patient Inclusion/Exclusion criteria and final cohort^2^

### 2.3. Study Measures

The available baseline and pre-index patient data were utilized to train the ML algorithms for the prediction of buprenorphine discontinuation. Treatment discontinuation was defined as a gap of ≥30 days without a buprenorphine prescription within one year of treatment initiation (Meinhofer, 2019; Samples, 2018). During the follow-up period, we measured medication adherence in the early phase of treatment by using the proportion of days covered (PDC) as a predictor. The PDC is calculated by dividing the number of days a patient had the medication covered by the total number of days in a specific timeframe. We calculated one-month and three-month PDC for each patient in the study and measured them as continuous variables.

### 2.4. Predictor candidates

Our predictor candidates (n=33), including patients’ socio-demographics, clinical condition and health status factors, are based on prior studies of buprenorphine therapy and medication adherence for MOUD (Lo-Ciganic, 2016; Williams, 2020; Hasan M. M., 2021; Stafford, 2022). Sociodemographic predictors included age, sex, geographic region (Northeast, Northcentral, South, West, Unknown), insurance plan type (EPO/PPO^3^, HMO/POS^4^, Others), relationship to policyholder (Employee, Spouse, Child/Other), and rural versus urban residence. The inclusion of these predictors helps identify demographic and contextual influences on buprenorphine discontinuation rates, address potential disparities or area-specific challenges, and highlight the effect of comprehensive coverage. Clinical predictors included pre-index comorbid conditions and substance use severity level. Presence of additional medical conditions before the index date can provide valuable information about the overall health status of the patients. Also, patients with previous substance use diagnoses may face additional challenges during buprenorphine treatment that may help explore the relationship between substance use severity and treatment outcome (Garfield, 2010; Lo-Ciganic, 2016).

We included 10 comorbid conditions capturing prior history of clinical conditions, mental health, and substance use disorders. We also included Charlson Comorbidity Index (CCI), days supply of the first buprenorphine fill for each patient (Roffman, 2016; Edlund, 2014). We also created a series of binary variables (yes/no) of medication use during the baseline period including antidepressants, antipsychotics, mood stabilizers, benzodiazepines, nonbenzodiazepine stimulants, and opioid analgesics. Additionally, 2 substance use severity levels (mild or moderate/severe), defined by Diagnostic and Statistical Manual of Mental Disorders (DSM), were used for OUD and cannabis use disorder (CUD) (Foundation, 2021). International Classification of Diseases-10th Revision (ICD-10) codes provided in the Appendix A (Table A4) were used to identify OUD and comorbid conditions. We identified prescription drugs using RedBook^®^ to match drug names with NDCs listed on prescription claims (IBM, Red Book, 2021). The utilization of health services (inpatient visit, outpatient visit, and emergency department visit) was also examined in the analysis. As noted, to assess treatment adherence during the early follow-up phase (i.e., first one and three months), we calculated the PDC, which represents the percentage of days that a patient had buprenorphine medication available within a specified timeframe. We measured PDC within the first one month and three months after treatment initiation, presenting the actual PDC values separately.

A comprehensive description of all the predictors used for prediction is provided in Appendix A (Table A1). To perform sensitivity analysis, we classified the PDC into low or high adherence categories using a threshold of 80% or higher. This approach aligns with previous studies examining buprenorphine treatment adherence during the early treatment phase (Tkacz, 2014; Ronquest, 2018; Hasan M. M., 2021).

### 2.5. Machine-learning approaches and prediction performance evaluation

We randomly divided the cohort into training (developing algorithms) and testing (algorithm’s prediction performance evaluation) samples. Training (80%) and testing (20%) cohorts were randomly generated (4:1 ratio) with similar proportion of buprenorphine discontinuation in both samples. We used six different ML algorithms: multivariable logistic regression (LR), decision tree classifier (DT), random forests (RF), extreme gradient boosting (XGB), adaptive boosting (AdaBoost), and the ensemble of RF and XGB (RF-XGB). As noted before, we applied these algorithms to predict treatment discontinuation in two distinct treatment stages: (1) at the time of initiating treatment, and (2) after one month and three months following treatment initiation (Hasan M. M., 2021). The performance evaluation of the algorithms was conducted using the scikit-learn machine learning library in the python 3.10 software. Appendix A (Methods) describes the details for each of the machine learning approaches we used. Previous studies showed that these methods consistently yield good prediction results (Chu, 2008; Lo-Ciganic W. H., 2019; Hasan M. M., 2021). To perform a more robust analysis and improve predictive performance, the hyperparameters of the models were optimized using grid search and randomized search methods, based on the area under the receiver operating characteristic curve (AUROC). The parameters that were fine-tuned are included in Appendix A (Table A2) for corresponding models. We used recursive feature elimination with a cross-validation (RFECV) approach for automatic feature selection and dimensionality reduction (Mustaqim, 2021). RFECV starts with all features and gradually eliminates less important features in iterative fashion based on model performance. It evaluates the impact of the features’ removals using cross-validation, ensuring an unbiased assessment (Faysal, 2022).

To evaluate how well the model distinguishes between high-risk and low-risk patients for treatment discontinuation, we analyzed the discrimination performance using the test sample. This involved comparing the C-statistics of different models. We also compared the precision-recall curves across different methods using the same test sample. To enhance the practical value in a clinical setting and explain the ML models, we reported the top 20 most important predictors, providing valuable insights into the relevant variables for prediction. These predictors are generated using the SHapley Additive exPlanations (SHAP) analysis for comprehensive model explanation and interpretation (Nohara, 2019). SHAP measures the impact of each feature on predictions, considering interactions with other features and helps identify influential features by ranking them based on their importance. For precise identification of individuals at varying levels of risk, we conducted risk stratification by decile risk subgrouping. We used decile risk scores to stratify patients in the test sample based on their predicted probability of discontinuing buprenorphine treatment (Lo-Ciganic W. H., 2020). Within the top decile, we partitioned patients into three distinct strata, specifically targeting the upper percentiles (1^st^, 2^nd^ to 5^th^, and 6^th^ to 10^th^), to facilitate a closer examination of those individuals at the highest risk of treatment discontinuation. This approach provides a more detailed analysis of high-risk patients within the cohort, enhancing the rigor of the proposed framework. Additionally, we generated a calibration curve that allows us to compare the predicted probabilities generated by our model with the actual probabilities observed in the dataset. The calibration curve is a graphical representation that assesses the calibration performance of a predictive model (Austin, 2020). We applied the isotonic calibration method to adjust predicted probabilities and observed an improvement in calibration. The overall accuracy of probabilistic predictions was assessed using the Brier score metric, where a lower score indicates better agreement between the predicted probabilities and the observed outcomes (Fenlon, 2018). When the probabilities increase and the curve moves upwards towards the top left, it indicates that for higher probabilities, the model predicts a particular outcome with high certainty, but the actual observed frequency of those outcomes is less than the model’s predicted probability (Niculescu-Mizil, 2005; Austin, 2020). This analysis serves as a crucial measure to evaluate the calibration of our model and determine if the predicted probabilities align well with the observed outcomes. All the analyses were performed and evaluated in two stages: at the time of initiating treatment, and after one month and three months following treatment initiation. It ensures that the model’s reliability and accuracy were robustly tested, emphasizing the strength of our proposed framework.

## 3. Results

### 3.1. Patient characteristics

The study sample included 30,373 eligible patients (mean age 40.92±12.11 years and 43.15% female). Table 1 presents descriptive statistics for patient characteristics stratified by buprenorphine continuation/discontinuation status. During the one-year follow-up period, 4551 patients (14.98%) discontinued treatment. The patients who discontinued buprenorphine were more likely to be younger (e.g., mean age: 39.77±12.52 versus 41.13±12.03 at the one-month after treatment initiation) (p value<0.001) and had lower PDC (p value<0.001) during both of the initial one-month and three-month treatment periods compared to those who continued buprenorphine. The mean PDC was 92% and 82% within the first one month and three months following treatment initiation, respectively.

**Table 1.** Characteristics of MarketScan commercially insured patients by buprenorphine discontinuation status during the first-year treatment.

|  | Total | Discontinued | Continued | Unadjusted RR<sup>5</sup><br>(95% CI) | P value<sup>6</sup> |
| --- | --- | --- | --- | --- | --- |
| **Number of patients** | 30373 | 4551 | 25822 | - | - |
| **Age, mean (SD)** | 40.92 (12.11) | 39.77 (12.52) | 41.13 (12.03) | N/A | <0.001 |
| **Age group, n (%)** |  |  |  |  |  |
| 18-24 | 3177 (10.46) | 723 (15.89) | 2454 (9.50) | Reference | Reference |
| 25-34 | 6986 (23.00) | 1004 (22.06) | 5982 (23.17) | 0.63 (0.58 – 0.69) | <0.001 |
| 35-44 | 8476 (27.91) | 1172 (25.75) | 7304 (28.29) | 0.61 (0.56 – 0.66) | <0.001 |
| 45-54 | 6383 (21.02) | 907 (19.93) | 5476 (21.21) | 0.62 (0.57 – 0.68) | <0.001 |
| 55-64 | 5351 (17.62) | 745 (16.37) | 4606 (17.84) | 0.61 (0.56 – 0.67) | <0.001 |
| **Sex, n (%)** |  |  |  |  |  |
| Male | 17267 (56.85) | 2605 (57.24) | 14662 (56.78) | 1.02 (0.96 – 1.07) | 0.56 |
| Female | 13106 (43.15) | 1946 (42.76) | 11160 (43.22) | Reference | Reference |
| **Geographic region, n (%)** |  |  |  |  |  |
| Northeast | 5022 (16.53) | 749 (16.46) | 4273 (16.55) | Reference | Reference |
| Northcentral | 5319 (17.51) | 713 (15.67) | 4606 (17.84) | 0.90 (0.82 – 0.99) | 0.03 |
| South | 15313 (50.42) | 2303 (50.60) | 13010 (50.38) | 1.01 (0.93 – 1.09) | 0.83 |
| West | 4658 (15.34) | 775 (17.03) | 3883 (15.04) | 1.12 (1.02 – 1.22) | 0.02 |
| Unknown | 61 (0.20) | 11 (0.24) | 50 (0.19) | 1.21 (0.71 – 2.07) | 0.50 |
| **Urban/Rural residence, n (%)** |  |  |  |  |  |
| Urban | 22033 (72.54) | 3359 (73.81) | 18674 (72.32) | Reference | Reference |
| Rural | 4766 (15.69) | 657 (14.44) | 4109 (15.91) | 0.90 (0.84 – 0.98) | 0.01 |
| Unknown/Missing | 3574 (11.77) | 535 (11.76) | 3039 (11.77) | 0.98 (0.90 – 1.07) | 0.67 |
| **Insurance plan type, n (%)** |  |  |  |  |  |

<sup>5</sup> Abbreviation: RR: Risk Ratio.
<sup>6</sup> P values were derived from a chi-square test for categorical variables and a t-test for continuous variables.

|  |  |  |  |  |  |
| --- | --- | --- | --- | --- | --- |
| EPO/PPO<sup>7</sup> | 16462<br>(54.20) | 2360 (51.86) | 14102<br>(54.61) | 0.88 (0.83 – 0.94) | <0.001 |
| HMO/POS<sup>8</sup> | 6280 (20.68) | 952 (20.92) | 5328 (20.63) | 0.93 (0.86 – 1.01) | 0.08 |
| Others | 7631 (25.12) | 1239 (27.22) | 6392 (24.75) | Reference | Reference |
| **Relationship to policyholder, n (%)** |  |  |  |  |  |
| Employee | 17214<br>(56.68) | 2464 (54.14) | 14750<br>(57.12) | 0.71 (0.66 – 0.77) | <0.001 |
| Spouse | 9237 (30.41) | 1299 (28.54) | 7938 (30.74) | 0.70 (0.65 – 0.76) | <0.001 |
| Child/Other | 3922 (12.91) | 788 (17.31) | 3134 (12.14) | Reference | Reference |
| **Comorbidities, n (%)** |  |  |  |  |  |
| **Charlson Comorbidity Index, mean (SD)** | 0.34 (0.96) | 0.31 (0.87) | 0.34 (0.97) | N/A | 0.05 |
| **Individual medical conditions** |  |  |  |  |  |
| Chronic pain | 8113 (26.71) | 1085 (23.84) | 7028 (27.22) | 0.81 (0.76-0.86) | <0.001 |
| HIV/AIDS | 128 (0.42) | 30 (0.66) | 98 (0.38) | 1.42 (1.04-1.94) | 0.04 |
| Hepatitis C | 3278 (10.79) | 515 (11.32) | 2763 (10.70) | 0.95 (0.87-1.04) | 0.25 |
| **Mental health disorders** |  |  |  |  |  |
| Depressive disorder | 4905 (16.15) | 740 (16.26) | 4165 (16.13) | 1.01 (0.94-1.09) | 0.76 |
| Anxiety | 5295 (17.43) | 813 (17.86) | 4482 (17.36) | 1.03 (0.96 – 1.10) | 0.43 |
| PTSD<sup>9</sup> | 610 (2.01) | 97 (2.13) | 513 (1.99) | 1.07 (0.89 – 1.28) | 0.50 |
| Bipolar disorder | 1169 (3.85) | 173 (3.80) | 996 (3.86) | 0.99 (0.86 – 1.14) | 0.91 |
| Schizophrenia | 133 (0.44) | 26 (0.57) | 107 (0.41) | 1.31 (0.93 – 1.85) | 0.14 |
| **Substance use disorders** |  |  |  |  |  |
| Non-opioid drug use disorder | 5676 (18.69) | 913 (20.06) | 4763 (18.45) | 1.09 (1.02-1.17) | 0.01 |
| Alcohol use disorder | 2012 (6.62) | 354 (7.78) | 1658 (6.42) | 1.20 (1.08-1.32) | <0.001 |
| **Substance use severity level, n (%)** |  |  |  |  |  |
| **Opioid use disorder (OUD)** |  |  |  |  |  |
| Mild | 979 (3.22) | 178 (3.91) | 801 (3.10) | 1.24 (1.08-1.43) | 0.002 |
| Moderate/Severe | 11000<br>(36.22) | 1681 (36.94) | 9319 (36.09) | 1.04 (0.99-1.10) | 0.13 |
| **Cannabis use disorder (CUD)** |  |  |  |  |  |
| Mild | 296 (0.97) | 42 (0.92) | 254 (0.98) | 0.95 (0.72-1.26) | 0.73 |
| Moderate/Severe | 612 (2.01) | 119 (2.61) | 493 (1.91) | 1.31 (1.11-1.54) | <0.001 |
| **Health services, n (%)** |  |  |  |  |  |
| Inpatient Visits | 4597 (15.14) | 821 (18.04) | 3776 (14.62) | 1.21 (1.11-1.31) | <0.001 |

<sup>7</sup> EPO: Exclusive Provider Organization Plan; PPO: Preferred Provider Organization Plan.
<sup>8</sup> HMO: Health Maintenance Organization Plan; POS: Point-of-Service Plan.
<sup>9</sup> PTSD: Post-traumatic stress disorder.

|  |  |  |  |  |  |
| --- | --- | --- | --- | --- | --- |
| ED Visits<sup>10</sup> | 6657 (21.92) | 1084 (23.82) | 5573 (21.58) | 1.10 (1.02-1.19) | 0.02 |
| Outpatient Visits | 22262<br>(73.30) | 3302 (72.56) | 18960<br>(73.43) | 1.00 (0.94-1.07) | 0.96 |
| **Pre-index medication use, n (%)** |  |  |  |  |  |
| Antidepressants | 11914<br>(39.23) | 1782 (39.16) | 10132<br>(39.24) | 1.03 (0.97-1.10) | 0.27 |
| Antipsychotics | 3049 (10.04) | 518 (11.38) | 2531 (9.80) | 1.18 (1.08-1.28) | <0.001 |
| Mood stabilizers | 7122 (23.45) | 1061 (23.31) | 6061 (23.47) | 1.03 (0.96-1.10) | 0.40 |
| Benzodiazepines | 6373 (20.98) | 995 (21.86) | 5378 (20.83) | 1.08 (1.01-1.16) | 0.03 |
| Nonbenzodiazepines | 1812 (5.97) | 256 (5.63) | 1556 (6.03) | 0.98 (0.87-1.10) | 0.71 |
| Stimulants | 2681 (8.83) | 473 (10.39) | 2208 (8.55) | 1.22 (1.11-1.34) | <0.001 |
| Opioid analgesics | 4184 (13.78) | 531 (11.67) | 3653 (14.15) | 0.88 (0.80-0.96) | 0.004 |
| **Initial days supply**<br>**for buprenorphine,**<br>**mean (SD)** | 19.93 (11.04) | 19.58 (11.14) | 20.00 (11.02) | N/A | 0.01 |
| **Early buprenorphine adherence, mean (SD)** |  |  |  |  |  |
| One-month PDC | 0.92 (0.20) | 0.87 (0.26) | 0.93 (0.19) | N/A | <0.001 |
| Three-month PDC | 0.82 (0.28) | 0.69 (0.31) | 0.84 (0.27) | N/A | <0.001 |
| **PDC within first month, n (%)** |  |  |  |  |  |
| High (≥ 80%) | 26686<br>(87.86) | 3617 (79.48) | 23069<br>(89.34) | Reference | Referenc<br>e |
| Low (< 80%) | 3687 (12.14) | 934 (20.52) | 2753 (10.66) | 1.87 (1.75 – 1.99) | <0.001 |
| **PDC within first three months, n (%)** |  |  |  |  |  |
| High (≥ 80%) | 20700<br>(68.15) | 2086 (45.84) | 18614<br>(72.09) | Reference | Referenc<br>e |
| Low (< 80%) | 9673 (31.85) | 2465 (54.16) | 7208 (27.91) | 2.53 (2.40 – 2.67) | <0.001 |

Table 1 also provides the unadjusted risk ratios (RR) and corresponding p-values for the risk of treatment discontinuation that we computed for different patient characteristics. Patients whose insurance plan type was PPO (RR=0.88, 95% confidence interval (CI) = 0.83-0.94, p <0.001) was associated with a decreased risk for treatment discontinuation compared to those who had HMO or other insurance. Patients having a mild level of pre-index OUD had a higher risk of discontinuation (RR=1.24, 95% CI= 1.08-1.43, p =0.002) compared to those without OUD. Also, patients having moderate or severe level of pre-index CUD were associated with a higher risk of discontinuation (RR=1.31, 95% CI= 1.11-1.54, p value<0.001) compared to those without CUD. In terms of health service utilization in 6-month pre-index period, the majority of the patients in the study sample had at least one encounter in an outpatient setting. However, patients with inpatient visits had a higher risk of discontinuation (RR=1.21, 95% CI= 1.11-1.31, p value<0.001) compared to those who had no inpatient visits. Similarly, patients with ED visits also showed an increased risk of discontinuation (RR=1.10, 95% CI= 1.02-1.19, p value=0.02) compared to those without ED visits. Among those with high PDC (≥80%) during the first one month and three months following treatment initiation, 13.55% and 10.08% discontinued treatment, respectively. By contrast, over 25% of the patients who had low PDC (<80%) in the first 1 month and 3 months of treatment initiation discontinued treatment. Patients with low PDC were associated with a significantly increased risks of buprenorphine discontinuation (the first month after initiation: RR=1.87, 95% CI=1.75-1.99; the first 3 months after initiation: RR=2.53, 95% CI=2.40-2.67, both p values<0.001) compared to those with high PDC.

### 3.2. Prediction performance across machine-learning models

Table A3 (shown in Appendix A) summarizes the discriminative and predictive performance (C-statistics and recall) of six ML models that we applied in two treatment stages. Table A3 also shows a comparison of the performance of these models before and after we implemented the RFECV technique. The findings revealed that performance of the models was moderately enhanced through the implementation of hyperparameter optimization and feature selection technique (i.e., RFECV). Figure 3 (left panels) graphically presents the models’ AUROC (C-statistics) values that we obtained after eliminating less important predictors with RFECV. The initial evaluation with the baseline predictors achieved the following C-statistics (Figure 3A): LR (C-statistics = 0.56), DT (0.58), RF (0.76), XGB (0.61), AdaBoost (0.58), and ensemble of RF-XGB (0.70). After incorporating the PDC measure of one-month from treatment initiation, the C-statistics improved to (Figure 3B): LR (0.60), DT (0.62), RF (0.79), XGB (0.65), AdaBoost (0.63), and ensemble of RF-XGB (0.73). As we further enhanced the models by including 3-month PDC, the overall trend of increasing C-statistics continued across all predictive models (Panel C): LR (0.65), DT (0.67), RF (0.82), XGB (0.71), AdaBoost (0.68), and ensemble of RF-XGB (0.77). RF outperformed all other models in predictive performance, with the highest C-statistic in all three cases. Recall values for different ML models after using RFECV ranged from 0.56 to 0.64 for baseline predictors; 0.60 to 0.65 after the inclusion of one-month PDC and 0.60 to 0.67 with three-month PDC. The models exhibited moderate improvements in their C-statistics when including information about patients’ medication adherence during the early treatment phases (i.e., one-month and three-month PDC). The precision-recall curve after using RFECV is also shown for all settings in Figure 3 (right panels).

**Figure 3.**
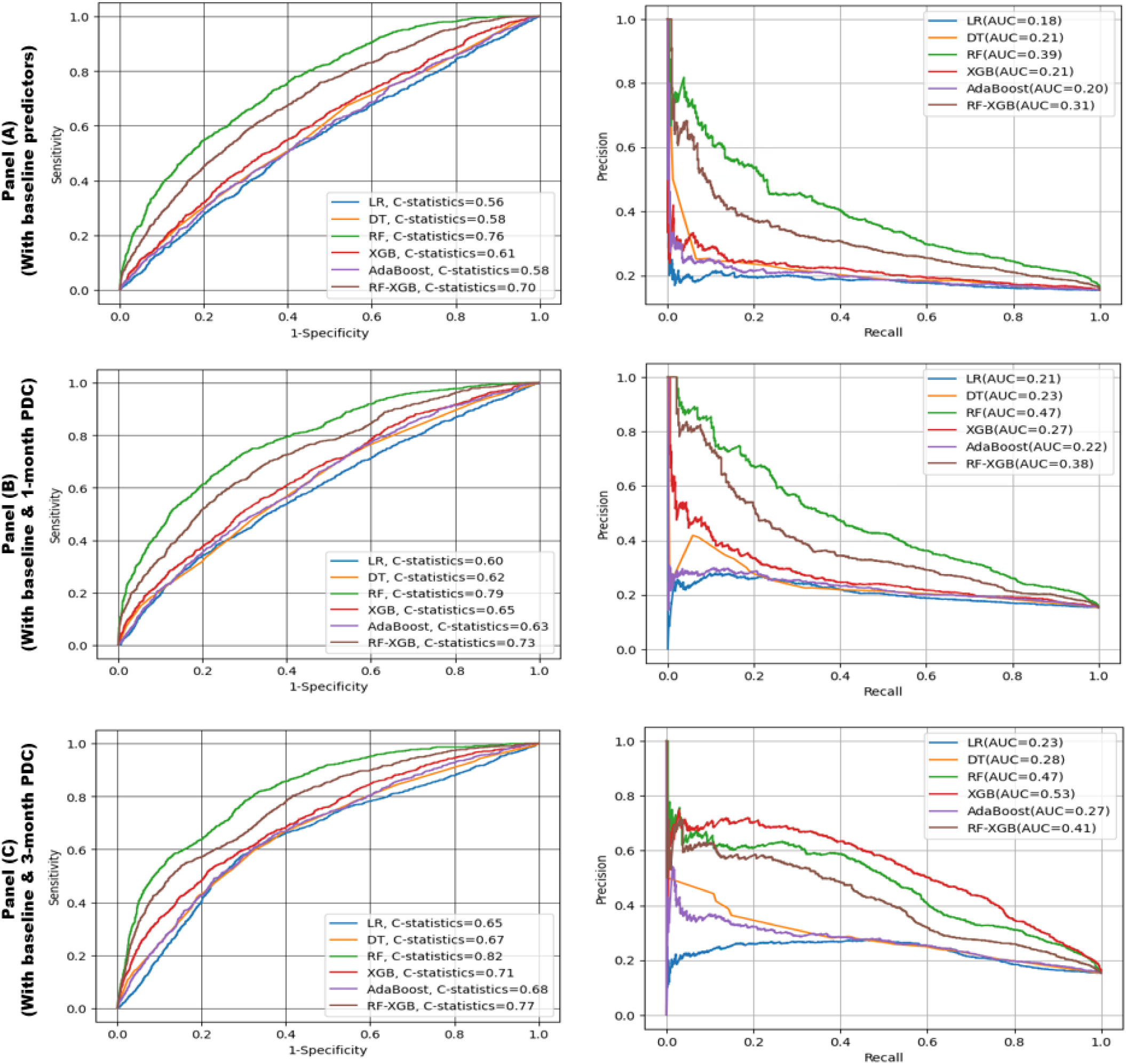
ROC curve^11^ after using RFECV for: 1st-stage model including baseline predictors (Panel A-left); 2nd-stage model including baseline predictors and one-month PDC as a continuous variable (Panel B-left); 2nd-stage model including baseline predictors and three-month PDC as a continuous variable (Panel C-left); Precision-Recall curve for: 1st-stage model including baseline predictors (Panel A-right); 2nd-stage model including baseline predictors and one-month PDC as a continuous variable (Panel B-right); 2nd-stage model including baseline predictors and three-month PDC as a continuous variable (Panel C-right). Precision-recall curves that are closer to the upper right corner or are above another method have improved performance.

### 3.3. Important predictors

As shown in Figure 4, we used SHAP analysis for model explanation and interpretation that provides a dual benefit. Figure 4 presents two variable importance plots utilizing SHAP values derived from the XGB model. The model incorporates the PDC measure as a continuous variable over one month (as depicted in Panel A) and three months (as illustrated in Panel B).

**Figure 4.**
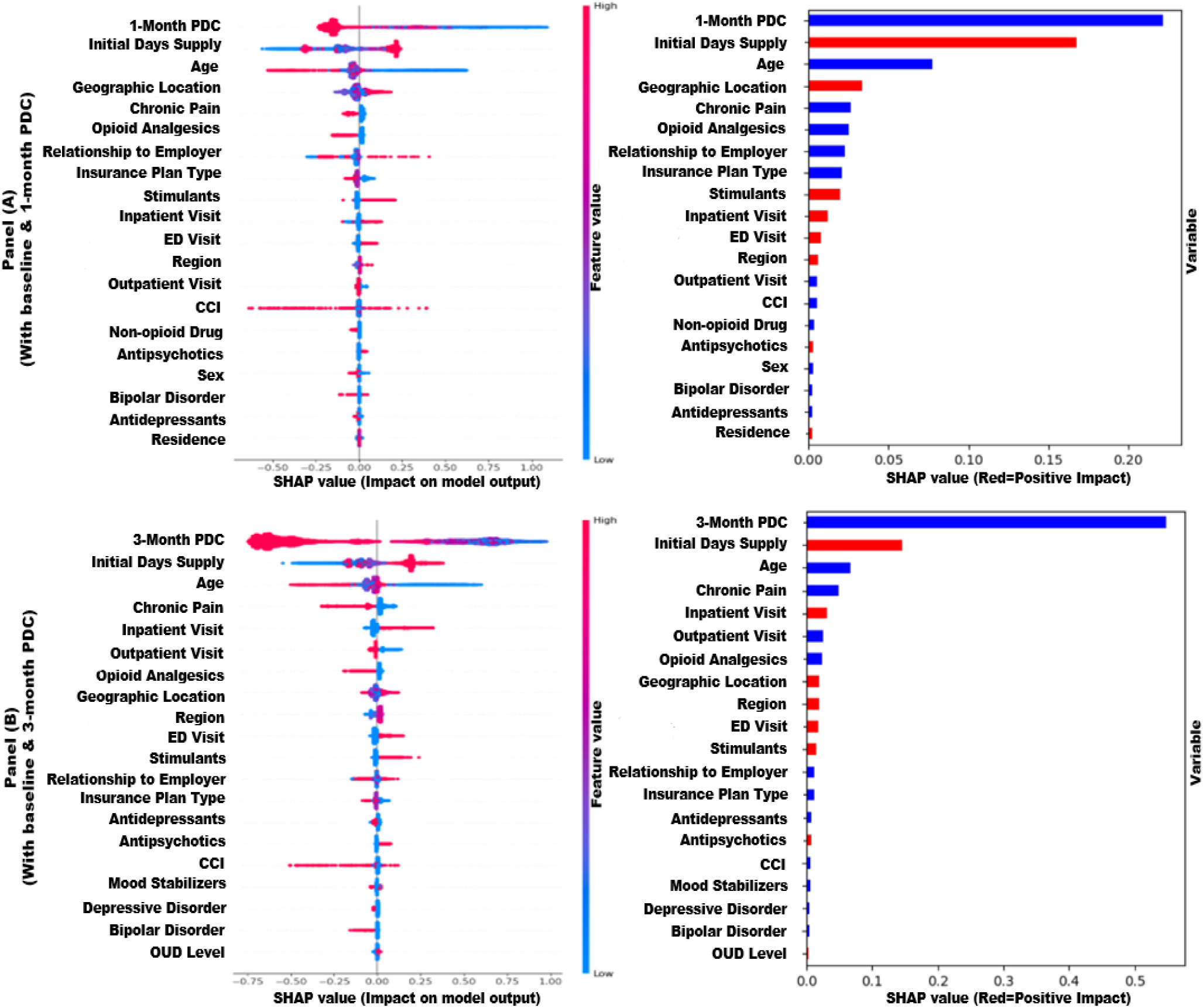
Variable importance plots using SHAP values from extreme gradient boosting (XGB) modeling including one-month PDC (Panel A) and three-month PDC (Panel B) as a continuous measure; SHAP value computed from individual feature’s values and their impact (both positive and negative) on treatment discontinuation (left in Panel A and Panel B); Average SHAP values of features showing average impact on and correlation with treatment discontinuation (right in Panel A and Panel B)

In Figure 4, the plots on the right-hand side of Panel A and Panel B present the average impact of various features on treatment discontinuation, listed in descending order of their significance. These plots indicate whether the mentioned features have a positive correlation (red color) or a negative correlation (blue color) with treatment discontinuation. The PDC value during the initial phase of treatment (in one month and three months) shows the most significant negative correlation with treatment discontinuation (e.g., higher PDC values associated with treatment continuation). Variables such as age, presence of chronic pain, use of opioid analgesics, type of insurance plan, and number of outpatient visits were negatively correlated with buprenorphine discontinuation, while factors having a positive correlation with treatment discontinuation included the initial days of supply of buprenorphine, the geographical location of the patient, number of inpatient visits, number of ED visits, and the use of stimulants.

### 3.4. Model calibration

To evaluate the model calibration, which measures the agreement between predicted and observed probability of discontinuation, we created calibration plots using the best performing RF (with three-month PDC) model (Figure 5). The initial RF model had a calibration curve situated in the upper region from the diagonal (perfectly calibrated) line, indicating poor calibration. After applying isotonic calibration, the curve moved closer to the diagonal line, signifying improved calibration. The Brier score metric used to measure the overall accuracy of probabilistic predictions, decreased from 0.17 to 0.12, indicating increased accuracy in the model’s predicted probabilities. This implies that the recalibrated model provides more reliable and accurate probability estimates for the discontinuation prediction. However, as the probabilities increased, the curve went upwards towards the top left indicating that the model can predict discontinuation with a high level of certainty, but the actual observed frequency of discontinuation is less than the model’s predicted probability.

**Figure 5.**
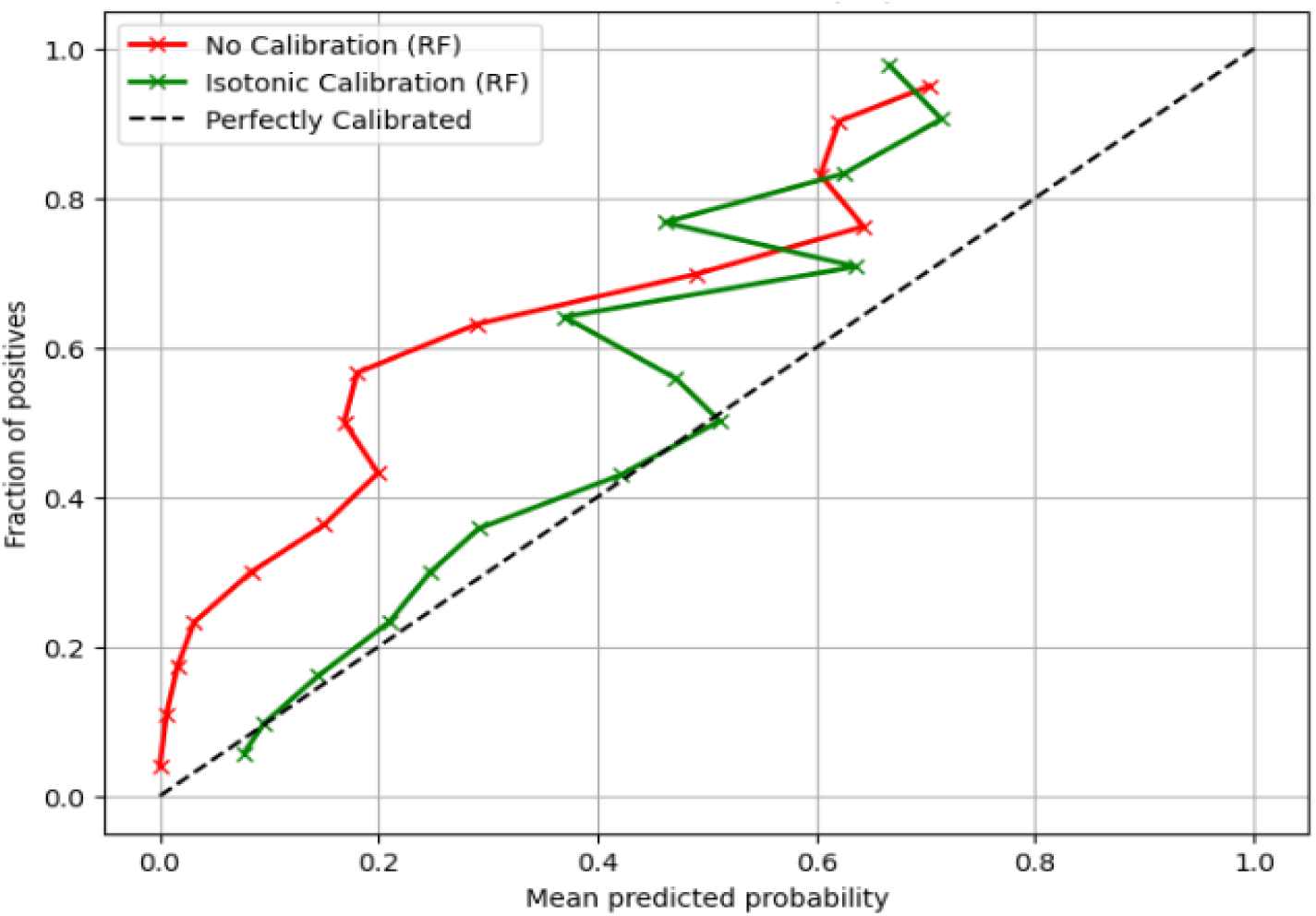
Calibration curve (with three-month PDC model) for RF

### 3.5. Risk stratification by decile risk subgroup

Figure 6 presents test sample’s patient subgroups based on their predicted probability of discontinuing OUD treatment where the percentile cutoff was derived from the training sample. The highest-risk subgroup (risk scores in the top 1^st^ percentile; 1% [n=61]) had a positive predictive value of 60.66%. The second highest-risk subgroup, with a risk score of 4% (n=243), had a positive predictive value of 41.98%. This was followed by the third highest-risk subgroup, which had a risk score of 5% (n=304) and a positive predictive value of 26.64%. Among 934 individuals with treatment discontinuation in the test set, 501 (53.6%) individuals were in the top three deciles of risk scores. Those in the top 1^st^ percentile had an 8-fold higher buprenorphine discontinuation rate compared to the lower-risk groups (e.g., observed discontinuation rate: top 1^st^ percentile of Decile 1 = 60.66%, Decile 10 = 7.25%).

**Figure 6.**
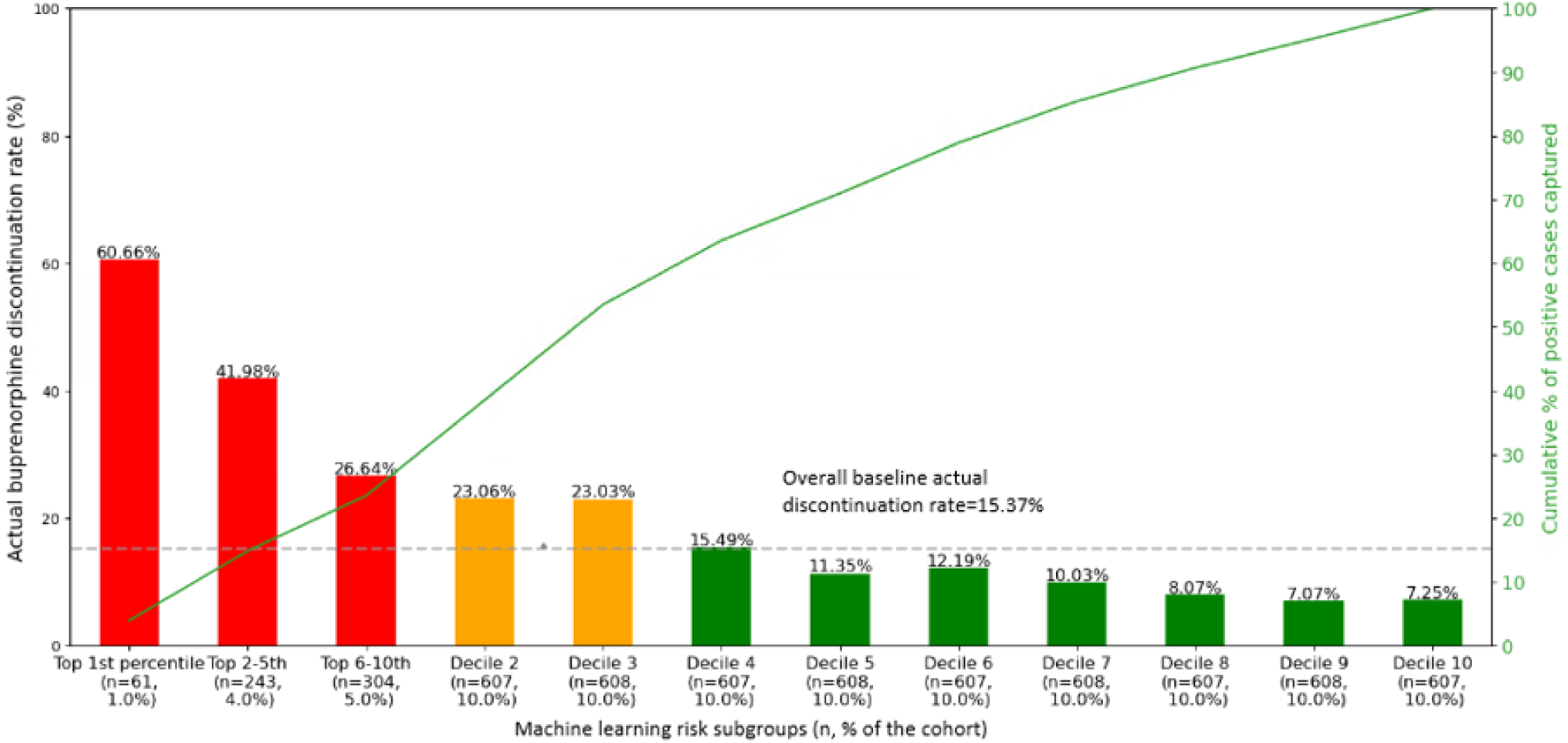
OUD^12^ treatment discontinuation identified by random forest’s decile risk subgroup in the test sample (n = 6075)

## 4. Discussion

This study represents a significant advancement on predicting buprenorphine treatment discontinuation, addressing the gaps in previous work and presenting a more robust analysis conducted using recent-years data from a large, nationally representative dataset. Our study’s robust two-stage clinical decision support framework utilized machine learning to predict patients’ buprenorphine discontinuation. This novel approach was designed with the goal of providing insights to improve patient adherence to buprenorphine and overall treatment outcomes. Our risk stratification approach that categorized patients into subgroups with similar risks of buprenorphine treatment discontinuation enhances our algorithm’s clinical utility and can efficiently direct interventions by tailoring them to patients’ specific risk profiles. The significant implications of our study’s results emphasize the importance of considering patient adherence information, optimizing model parameters, and utilizing feature selection/dimensionality reduction techniques to develop more robust and reliable predictive models. The insights derived from our findings can be leveraged in real-world clinical settings to potentially inform treatment strategies, thereby minimizing buprenorphine discontinuation and optimizing patient outcomes.

While our first-stage or baseline models did not yield a considerably high C-statistic value, they were improved when incorporating information about patients’ medication adherence measured over the early treatment phase. These findings significantly contribute to our understanding of the trajectory of patient adherence. It points to the critical role of early treatment adherence in the prediction of OUD treatment discontinuation. All the models achieved moderate to good C-statistics, with random forest outperforming other models in all three cases (baseline, inclusion of one-month PDC and three-month PDC). This finding indicates the superior discrimination ability of the state-of-the-art ML models, indicating their potential to be used in similar predictive scenarios.

This analysis of a recent, large, nationally representative dataset of commercially insured patients revealed that younger patients in general had a higher risk of discontinuation, a finding which has also been reported in previous studies with a sample of commercially insured beneficiaries from one state (i.e., Massachusetts), a national sample of individuals receiving outpatient specialty treatment, a multi-state sample of Medicaid beneficiaries, an older (2010-2014) sample of commercially insured, US beneficiaries (Hasan M. M.-E.-A., 2021; Krawczyk N. W., 2021; Samples, 2018; Morgan, 2018). This finding is indicative of the additional support that younger populations may require appropriate interventions to ensure sustain treatment. Further, the study also identified that the mean PDC during the early phase following treatment initiation was higher among patients who continued treatment compared to those who discontinued. This observation aligns with previous research (Hasan M. M., 2021), underlining the importance of medication adherence in early treatment stages to promote long-term treatment retention. Therefore, interventions focusing on enhancing adherence in the early phase of treatment could be crucial in improving patient outcomes.

The accuracy of our predictive models was further ascertained through model calibration that verified the reliability of the models. Our models demonstrated not only strong discriminative ability but also moderate calibration, indicating their potential to be reliable tools in predicting buprenorphine treatment discontinuation. This analytical approach allows us to gain a better understanding of our models’ prediction capabilities, highlighting their ability to predict outcomes that closely align with the actual observed data. Using risk stratification by decile risk subgroup, we precisely identified individuals across varying risk levels and gathered valuable insights about the characteristics of the patients who were at the highest risk of buprenorphine discontinuation. This risk stratification has the potential to enhance personalized treatment plans by providing targeted interventions for high-risk patients. These plans may include intensive monitoring, personalized therapy, medication adjustments, care coverage expansion, community involvement, etc. (Lee, 2018; Meyer, 2021). This combination of discriminative and predictive power, along with patient risk stratification capabilities, gives our models a distinct edge in clinical decision-making contexts.

Although the findings of the study are promising, some limitations need to be acknowledged. First, the application of predictive ML methods in this study yield findings that are associative rather than causative. This is an inherent limitation of using ML-based approaches and something that should be taken into consideration when interpreting the results. Second, we excluded patients who had <2 buprenorphine fills. This exclusion may have eliminated those who started their treatment in the ED setting, which is intended more for acute situations than for long-term care (Pines, 2011). Establishing continuity of care from EDs to outpatient settings often poses a challenge, particularly for individuals who lack stable housing or are involved in the justice system (Krawczyk N. B., 2019; Stoller, 2016). Future research is needed to study this ED population. Third, our study cohort included continuously enrolled patients throughout the observation period. This could be a potential limitation of our findings, as it may not directly apply to patients who lose their insurance or experience gaps in coverage, which are important factors contributing to OUD treatment discontinuity (Zeledon, 2020). Fourth, the study did not consider unmeasured confounders such as patient motivation and support systems, which might have influenced the results. Their inclusion in future analyses may provide a more holistic view of the patient’s treatment journey and reduce potential bias in predicting treatment discontinuation. Additionally, we could not see data for patients who paid in cash for MOUD treatment. However, by identifying key risk factors for buprenorphine treatment discontinuation among continuously insured patients, our study contributes valuable data and evidence. It allows us to understand the discontinuation patterns in a more controlled environment, where financial barriers to treatment are less likely to be the primary cause of discontinuation. This study can demonstrate the need to shift the current recommended set of targeted interventions away from being limited to siloed MOUD care linkages and getting back to root causes that are really about maintaining or linking to insurance coverage as a whole.

## 5. Conclusion

The findings of this study suggest that machine learning models can effectively identify insured high-risk patients for OUD treatment discontinuation, thereby providing a potentially valuable tool for helping healthcare professionals make more informed clinical decisions. The proposed framework is robust in terms of discriminative power, reliability and patients’ risk stratification. The systematic integration of ML algorithms, explainable methods, and patients’ risk stratification provides a unique approach for targeted interventions of OUD patients. This represents a significant step forward in the field of substance use disorder treatment, where the prediction and prevention of treatment discontinuation have always been a complex issue. Utilizing such frameworks for predicting treatment discontinuation may assist healthcare providers to design tailored support systems that enhance the long-term retention of patients in OUD treatment with buprenorphine.

## CRediT authorship contribution statement

**Jabed Al Faysal:** Methodology, Software, Formal analysis, Data curation, Visualization, Validation, Writing – original draft, editing. **Md. Noor-E-Alam:** Writing – review & editing, Supervision. **Gary J. Young:** Writing – review & editing, Supervision. **Wei-Hsuan Lo-Ciganic:** Writing – review & editing, Validation, Resources. **Amie J. Goodin:** Writing – review & editing, Validation. **James L. Huang:** Software, Visualization. **Debbie L. Wilson:** Writing – review & editing. **Tae Woo Park:** Writing – review. **Md Mahmudul Hasan:** Conceptualization, Investigation, Writing – review & editing, Validation, Resources, Supervision.

## Data Availability

All data produced in the present study are available upon reasonable request to the authors

## Acknowledgements

I would like to express my sincere gratitude to all authors who have contributed to the completion of this project.

## Footnotes

1 Abbreviation: PDC: Proportion of Days Covered.

2 Abbreviation: OUD: opioid use disorder. Definitions: buprenorphine discontinuation: having a gap of ≥30 days without a buprenorphine prescription within the first year of treatment initiation. buprenorphine continuation: having no gap of ≥30 days without a buprenorphine prescription within the first year of treatment initiation

3 EPO: Exclusive Provider Organization Plan; PPO: Preferred Provider Organization Plan.

4 HMO: Health Maintenance Organization Plan; POS: Point-of-Service Plan.

10 ED visit: Emergency Department visit.

11 ROC: Receiver Operating Characteristics; LR: Logistic Regression; DT: Decision Tree; RF: Random Forest; XGB: Extreme Gradient Boosting; AdaBoost: Adaptive Boosting; RF-XGB: Ensemble of Random Forest and Extreme Gradient Boosting; PDC: Proportion of Days Covered; RFECV: Recursive Feature Elimination with Cross-Validation.

12 OUD: opioid use disorder.

