## Appendix A for "An Explainable Machine Learning Framework for Predicting the Risk of Buprenorphine Treatment Discontinuation for Opioid Use Disorder"

### Methods: Machine Learning Algorithms, Feature Selection Technique and Explainable AI

**Multivariable Logistic Regression (LR)**: Logistic regression is a widely used classification algorithm that models the relationship between the target variable and input features using logistic functions. It estimates the probability of belonging to a certain class and can handle both continuous and categorical variables. It is interpretable and performs well in linearly separable data. LR uses a threshold to classify instances into binary classes based on the estimated probabilities. LR assumes a linear relationship between the log odds of the outcome and the predictor variables. It applies the sigmoid function to restrict the predicted probability between 0 and 1. Also, being a parametric method, logistic regression requires the absence of multicollinearity among predictors and needs a large sample size to achieve stable results (Boateng, 2019).

**Decision Tree (DT)**: Decision trees create a hierarchical structure by recursively splitting data based on input features. They are interpretable and can handle both categorical and numerical features. Decision trees capture non-linear relationships and interactions between features but are prone to overfitting, which can be mitigated by ensemble methods. In binary classification, decision trees classify instances by assigning them to the class that is most prevalent in the leaf node they reach. Decision Trees have the added advantage of requiring minimal data preprocessing - scaling or normalization isn't necessary. They also provide feature importance, indicating which variables are most impactful in prediction. However, decision trees can be sensitive to changes in the data, meaning a small alteration in input can lead to a drastically different tree structure (Priyam, 2013).

**Random Forest (RF)**: Random forest is an ensemble learning algorithm that constructs multiple decision trees and combines their predictions through voting or averaging. It leverages random feature selection and bootstrapping to create diverse trees, reducing overfitting. In binary classification, random forest assigns class labels based on the majority vote of the trees. It captures complex interactions between features and is effective in handling imbalanced datasets. Random Forests handle high dimensional spaces well and are resistant to outliers. The algorithm's inherent feature selection helps in dealing with collinearity. Despite their robustness and versatility, they do come with a downside–they lack the interpretability of a single decision tree, as the intuition behind the ensemble's decision can be hard to grasp (Biau, 2012).

**Extreme Gradient Boosting (XGB)**: XGB is a promising tree-based ensemble learning classifier, which is treated as the most effective implementation of gradient-boosted decision trees. Gradient boosted decision trees utilize a series of decision trees, each of which learns from the preceding tree and affects the subsequent tree; therefore, they improve the model and develop a powerful learner. It incorporates regularization techniques to prevent overfitting and supports various objective functions and evaluation metrics. In binary classification, XGBoost uses a threshold to classify instances based on the final boosted model's predicted scores. XGBoost also features in-built cross-validation, enabling the user to directly find the optimal number of boosting rounds. Furthermore, it's computationally efficient, with support for parallel processing, handling missing values, and tree-pruning, makes the model compact and faster to run (Ramraj, 2016).

**Adaptive Boosting (AdaBoost)**: AdaBoost is an iterative algorithm that combines multiple weak classifiers to create a strong classifier. It assigns higher weights to misclassified samples, allowing subsequent weak classifiers to focus on those samples and improve overall accuracy. In binary classification, AdaBoost assigns class labels based on the weighted votes of the weak classifiers. It adapts its weights at each iteration to emphasize the misclassified instances. AdaBoost is simple to implement and requires minimal configuration of hyperparameters, making it user-friendly. However, it is sensitive to noisy data and outliers, which can adversely impact performance (Feng, 2020).

**Ensemble of Random Forest and Extreme Gradient Boosting (RF-XGB)**: The ensemble model combines the predictions of RF and XGB, leveraging the strengths of both algorithms. By combining the diverse decision-making processes of random forest and the boosting power of XGB, this ensemble approach aims to improve prediction accuracy and generalization capabilities. In binary classification, the ensemble model can use voting or averaging of the predictions to determine the final class label for each instance. This hybrid model has an advantage of being robust against overfitting due to the random subspace method in RF, and also achieves higher predictive performance from XGB's gradient boosting (Faysal, 2022).

**Recursive Feature Elimination with Cross-Validation (RFECV)**: RFECV is an iterative process that starts with all features and gradually eliminates less important features based on model performance. It evaluates the impact of feature removal using cross-validation, ensuring unbiased assessment. RFECV specifically focuses on binary classification tasks. At each iteration, RFECV removes the least significant features and re-evaluates the model's performance. This recursive process continues until the desired number of features is reached or the optimal subset of features is determined. RFECV aims to select features that contribute the most to the distinction between the two binary classes. This method helps reduce dimensionality, improve model interpretability, thus enhancing the model's ability to discriminate between the two classes. The major difference between RFECV and RFE (Recursive Feature Elimination) is that in RFECV, the estimator is tested in terms of generating predictions on hold-out fold data in each feature subset. As a result, the best feature subset can be identified by ranking CV scores (Mustaqim, 2021).

**SHapley Additive exPlanations (SHAP)**: SHAP analysis, originated from cooperative game theory, is a technique for interpreting binary classification models by calculating Shapley values. It is a tool for Explainable AI (XAI) that focuses on making machine learning models more transparent and understandable to humans. SHAP measures the impact of each feature on predictions, considering interactions with other features and helps identify influential features by ranking them based on their importance. It provides local interpretations by explaining the contribution of each feature to individual predictions while it reveals overall feature importance across the entire dataset at the global level. SHAP aids in model validation, debugging, and addressing biases. It also enables feature interaction analysis, offering insights into how combinations of features affect predictions, enhancing model transparency and trustworthiness (Nohara, 2019).

### Predictor Descriptions

#### Table A1. Summary of predictor candidates for predicting opioid use disorder treatment discontinuation

| Predictor Category | Predictor Names | Description |
|---|---|---|
| **Demographic** | Age | Age of patient |
| | Sex | Gender of patient |
| | Geographic Region | Geographic region of the patient |
| | Residence | Urban/Rural residence |
| | Insurance Plan Type | Type of benefit plan |
| | Relationship to Policyholder | Relationship of the patient to the primary beneficiary |
| | Geographic location | Geographic location of the patient |
| **Health Status Factors** | Chronic pain | Chronic pain condition |
| | HIV/AIDS | HIV/AIDS condition |
| | Hepatitis C | Hepatitis C condition |
| | Depressive disorder | Depressive disorder condition |
| | Anxiety | Anxiety condition |
| | PTSD | Post-traumatic stress disorder condition |
| | Bipolar disorder | Bipolar disorder condition |
| | Schizophrenia | Schizophrenia condition |
| | Non-opioid drug use disorder | Non-opioid drug use condition |
| | Alcohol use disorder | Alcohol use condition |
| | Opioid use disorder (OUD) severity level | OUD severity level of the patient |
| | Cannabis use disorder (CUD) severity level | CUD severity level of the patient |
| | Charlson Comorbidity Index | Risk calculation with various comorbid conditions |
| | Initial Days Supply | No. of days in first buprenorphine prescription fill |
| **Medications Use** | Antidepressants | Use of Antidepressants |
| | Antipsychotics | Use of Antipsychotics |
| | Mood stabilizers | Use of Mood stabilizers |
| | Benzodiazepines | Use of Benzodiazepines |
| | Non-benzo | Use of Non-benzo |
| | Stimulants | Use of Stimulants |
| | Opioid analgesics | Use of opioid analgesics |
| **Health Service Utilization** | Inpatient Visits | No. of inpatient visits |
| | ED Visits | No. of emergency department (ED) visits |
| | Outpatient Visits | No. of outpatient visits |
| **Early Treatment Adherence** | 1-month PDC | Proportion of days covered (PDC) measure after 1-month of treatment initiation |

| | | |
|---|---|---|
| | 3-month PDC | Proportion of days covered (PDC) measure after 3-month of treatment initiation |

### Optimized Hyperparameters of the Models

#### Table A2. Machine learning model[1] hyperparameters in two-stages for the prediction of buprenorphine treatment discontinuation

| Treatment stage | ML model | Given hyperparameters | Selected hyperparameters |
|---|---|---|---|
| First stage models with baseline predictors | LR | solver: newton-cg, lbfgs, liblinear<br>penalty: l1, l2<br>penalty/regularization strength, C: 0.001, 0.01, 0.1, 1, 10 | solver: newton-cg<br>penalty: l2<br>C: 0.1 |
| | DT | criterion: gini, entropy<br>min_samples_leaf: 10, 20, 30, 40, 50 | criterion: gini<br>min_samples_leaf: 30 |
| | RF | n_estimators: 50,100,200<br>min_samples_leaf: 1,2,4<br>min_samples_split: 2,5,10<br>max_depth: 2,5,10 | n_estimators: 200<br>min_samples_leaf: 2<br>min_samples_split: 5<br>max_depth: 10 |
| | XGB | learning_rate: 0.01,0.05,0.1<br>n_estimators: 50,100,200<br>max_depth: 3,5,7<br>subsample: 0.5,0.75,1<br>colsample_bytree: 0.5,0.75,1 | learning_rate: 0.05<br>n_estimators: 100<br>max_depth: 3<br>subsample: 1<br>colsample_bytree: 0.75 |
| | AdaBoost | estimator: 1,2,3<br>N_estimators: 50,100,200<br>learning_rate: 0.01,0.05,0.1 | estimator: 1<br>N_estimators: 50<br>learning_rate: 0.1 |
| | RF-XGB | n_estimators (rf): 50,100,200<br>min_samples_leaf: 1,2,4<br>min_samples_split: 2,5,10<br>max_depth (xgb): none,5,10<br>learning_rate: 0.01,0.05,0.1<br>n_estimators (xgb): 50,100,200<br>max_depth (xgb): 3,5,7<br>subsample: 0.5,0.75,1<br>colsample_bytree: 0.5,0.75,1 | n_estimators (rf): 200<br>min_samples_leaf: 2<br>min_samples_split: 5<br>max_depth: 10<br>learning_rate: 0.05<br>n_estimators: 100<br>max_depth (xgb): 3<br>subsample: 1<br>colsample_bytree: 0.75 |
| Second stage models including 1-month PDC[2] as continuous measure | LR | solver: newton-cg, lbfgs, liblinear<br>penalty: l1, l2<br>penalty/regularization strength, C: 0.001, 0.01, 0.1, 1, 10 | solver: newton-cg<br>penalty: l2<br>C: 0.1 |
| | DT | criterion: gini, entropy<br>min_samples_leaf: 10, 20, 30, 40, 50 | criterion: gini<br>min_samples_leaf: 30 |
| | RF | n_estimators: 50,100,200<br>min_samples_leaf: 1,2,4<br>min_samples_split: 2,5,10<br>max_depth: 2,5,10 | n_estimators: 200<br>min_samples_leaf: 2<br>min_samples_split: 5<br>max_depth: 10 |
| | XGB | learning_rate: 0.01,0.05,0.1<br>n_estimators: 50,100,200<br>max_depth: 3,5,7 | learning_rate: 0.05<br>n_estimators: 100<br>max_depth: 3 |

[1] LR: Logistic Regression; DT: Decision Tree Classifier; RF: Random Forest; XGB: Extreme Gradient Boosting; AdaBoost: Adaptive Boosting; RF-XGB: Ensemble of Random Forest and Extreme Gradient Boosting.
[2] PDC: Proportion of Days Covered.

| | | | |
|---|---|---|---|
| | | subsample: 0.5,0.75,1<br>colsample_bytree: 0.5,0.75,1 | subsample: 1<br>colsample_bytree: 0.75 |
| | AdaBoost | estimator: 1,2,3<br>N_estimators: 50,100,200<br>learning_rate: 0.01,0.05,0.1 | estimator: 2<br>N_estimators: 100<br>learning_rate: 0.1 |
| | RF-XGB | n_estimators (rf): 50,100,200<br>min_samples_leaf: 1,2,4<br>min_samples_split: 2,5,10<br>max_depth (xgb): none,5,10<br>learning_rate: 0.01,0.05,0.1<br>n_estimators (xgb): 50,100,200<br>max_depth (xgb): 3,5,7<br>subsample: 0.5,0.75,1<br>colsample_bytree: 0.5,0.75,1 | n_estimators (rf): 200<br>min_samples_leaf: 2<br>min_samples_split: 5<br>max_depth: 10<br>learning_rate: 0.05<br>n_estimators: 100<br>max_depth (xgb): 3<br>subsample: 1<br>colsample_bytree: 0.75 |
| Second stage models including 3-month PDC as continuous measure | LR | solver: newton-cg, lbfgs, liblinear<br>penalty: l1, l2<br>penalty/regularization strength, C: 0.001, 0.01, 0.1, 1, 10 | solver: newton-cg<br>penalty: l2<br>C: 0.1 |
| | DT | criterion: gini, entropy<br>min_samples_leaf: 10, 20, 30, 40, 50 | criterion: gini<br>min_samples_leaf: 30 |
| | RF | n_estimators: 50,100,200<br>min_samples_leaf: 1,2,4<br>min_samples_split: 2,5,10<br>max_depth: 2,5,10 | n_estimators: 200<br>min_samples_leaf: 2<br>min_samples_split: 5<br>max_depth: 10 |
| | XGB | learning_rate: 0.01,0.05,0.1<br>n_estimators: 50,100,200<br>max_depth: 3,5,7<br>subsample: 0.5,0.75,1<br>colsample_bytree: 0.5,0.75,1 | learning_rate: 0.05<br>n_estimators: 100<br>max_depth: 3<br>subsample: 1<br>colsample_bytree: 0.75 |
| | AdaBoost | estimator: 1,2,3<br>N_estimators: 50,100,200<br>learning_rate: 0.01,0.05,0.1 | estimator: 2<br>N_estimators: 100<br>learning_rate: 0.1 |
| | RF-XGB | n_estimators (rf): 50,100,200<br>min_samples_leaf: 1,2,4<br>min_samples_split: 2,5,10<br>max_depth (xgb): none,5,10<br>learning_rate: 0.01,0.05,0.1<br>n_estimators (xgb): 50,100,200<br>max_depth (xgb): 3,5,7<br>subsample: 0.5,0.75,1<br>colsample_bytree: 0.5,0.75,1 | n_estimators (rf): 200<br>min_samples_leaf: 2<br>min_samples_split: 5<br>max_depth: 10<br>learning_rate: 0.05<br>n_estimators: 100<br>max_depth (xgb): 3<br>subsample: 1<br>colsample_bytree: 0.75 |

### Comparison of Machine Learning Model Performance in Different Settings

#### Table A3. Comparison of machine learning model[3] performance in different settings

| Before using RFECV[4] | | | After using RFECV | | |
|---|---|---|---|---|---|
| With baseline predictors | With baseline predictors & 1-month PDC[5] | With baseline predictors & 3-month PDC | With baseline predictors | With baseline predictors & 1-month PDC | With baseline predictors & 3-month PDC |
| **LR**<br>C-statistic: 0.55<br>Recall: 0.54<br>**DT**<br>C-statistic: 0.54<br>Recall: 0.55<br>**RF**<br>C-statistic: 0.66<br>Recall: 0.54<br>**XGB**<br>C-statistic: 0.57<br>Recall: 0.53<br>**AdaBoost**<br>C-statistic: 0.53<br>Recall: 0.49<br>**RF-XGB**<br>C-statistic: 0.62<br>Recall: 0.54 | **LR**<br>C-statistic: 0.60<br>Recall: 0.59<br>**DT**<br>C-statistic: 0.62<br>Recall: 0.58<br>**RF**<br>C-statistic: 0.69<br>Recall: 0.57<br>**XGB**<br>C-statistic: 0.63<br>Recall: 0.60<br>**AdaBoost**<br>C-statistic: 0.57<br>Recall: 0.51<br>**RF-XGB**<br>C-statistic: 0.66<br>Recall: 0.59 | **LR**<br>C-statistic: 0.64<br>Recall: 0.59<br>**DT**<br>C-statistic: 0.67<br>Recall: 0.59<br>**RF**<br>C-statistic: 0.70<br>Recall: 0.59<br>**XGB**<br>C-statistic: 0.69<br>Recall: 0.60<br>**AdaBoost**<br>C-statistic: 0.58<br>Recall: 0.51<br>**RF-XGB**<br>C-statistic: 0.70<br>Recall: 0.60 | **LR**<br>C-statistic: 0.56<br>Recall: 0.60<br>**DT**<br>C-statistic: 0.58<br>Recall: 0.64<br>**RF**<br>C-statistic: 0.76<br>Recall: 0.56<br>**XGB**<br>C-statistic: 0.61<br>Recall: 0.59<br>**AdaBoost**<br>C-statistic: 0.58<br>Recall: 0.64<br>**RF-XGB**<br>C-statistic: 0.70<br>Recall: 0.61 | **LR**<br>C-statistic: 0.60<br>Recall: 0.60<br>**DT**<br>C-statistic: 0.62<br>Recall: 0.60<br>**RF**<br>C-statistic: 0.79<br>Recall: 0.65<br>**XGB**<br>C-statistic: 0.65<br>Recall: 0.64<br>**AdaBoost**<br>C-statistic: 0.63<br>Recall: 0.60<br>**RF-XGB**<br>C-statistic: 0.73<br>Recall: 0.63 | **LR**<br>C-statistic: 0.65<br>Recall: 0.67<br>**DT**<br>C-statistic: 0.67<br>Recall: 0.60<br>**RF**<br>C-statistic: 0.82<br>Recall: 0.64<br>**XGB**<br>C-statistic: 0.71<br>Recall: 0.62<br>**AdaBoost**<br>C-statistic: 0.68<br>Recall: 0.60<br>**RF-XGB**<br>C-statistic: 0.77<br>Recall: 0.63 |

**Abbreviations:** RFECV = Recursive Feature Elimination with Cross-Validation; LR=Logistic Regression; DT=Decision Tree; RF=Random Forest; XGB=eXtreme Gradient Boosting, AdaBoost=Adaptive Boosting; RF-XGB=Ensemble of Random Forest and eXtreme Gradient Boosting; PDC=Proportion of Days Covered.

[3] Abbreviations: LR=Logistic Regression; DT=Decision Tree; RF=Random Forest; XGB=Extreme Gradient Boosting, AdaBoost=Adaptive Boosting; RF-XGB=Ensemble of Random Forest and Extreme Gradient Boosting.
[4] RFECV: Recursive Feature Elimination with Cross-Validation.
[5] PDC=Proportion of Days Covered.

### Health Status Factors and Codes

### Table A4. Diagnosis codes for identifying medical and mental health conditions

| Conditions | ICD-10 codes |
|---|---|
| Chronic pain | E0842, E0942, E1042, E1142, E1342, G43-G44, G50.1, G56.0, G56.4, G57, G58.9, G60-G65, G89.0, G89.2, G89.4, G90.0, G99.0, H46-H47, M00-M02, M05-M08, M11-M25, M30-M99, R26.2, R29.4, R29.898, R51 |
| HIV/AIDS | B20-B24 |
| Hepatitis C | B17.10, B17.11, B18.2, B19.20, B19.21 |
| Depressive disorder | F32, F33, F34.1 |
| Anxiety | F40-F42 |
| PTSD[6] | F43.1 |
| Bipolar disorder | F31, F34.0 |
| Schizophrenia | F20–F21, F25 |
| Non-opioid drug use disorder | F12–F19 |
| Alcohol use disorder | F10 |
